# Challenges and Opportunities Experienced by Performing Artists during COVID-19 Lockdown: Scoping Review

**DOI:** 10.1101/2022.02.23.22271390

**Authors:** Samantha K Brooks, Sonny S Patel

## Abstract

This scoping review synthesises published literature on the experiences of professional and amateur performing artists during COVID-19 and their perceptions of the challenges and opportunities faced. Six electronic databases were searched for published English-language articles containing primary data on this topic; twenty-one studies were reviewed. Themes included loss of work, financial impact, concerns about the future, psychological wellbeing, social connections, continuing creative pursuits, and inequalities. Participants reported both detrimental psychological effects of lockdown such as anxiety and sleep problems and positive effects including reduced stress and enjoyment of having more free time. Most continued creative pursuits throughout lockdown, most commonly shifting to online platforms. However, many barriers to creative pursuits were reported, including lack of technological expertise or equipment. Concerns were raised about inequality, in particular racial disparities in the financial impact of the pandemic and additional pressures faced by performers with disabilities; with insufficient funds to afford the equipment needed to shift to remote performing; and with additional caring responsibilities. It is important that performing artists have access to peer support; that education on digital technologies is incorporated into future performing arts education; and that inequities are addressed to ensure the needs of diverse communities are met.

## 1. Introduction

On March 11^th^ 2020, COVID-19 was declared to be a global pandemic [1]. Across the world, governments took action to try to reduce the spread of the virus, including closing all businesses deemed to be non-essential and imposing restrictions on socialising. Across all sectors, the pandemic disrupted working conditions: by April 2020, around 81% of the world’s workforce was reported to be affected by full or partial lockdown measures [2]. The lockdowns imposed due to COVID-19 have led to a global increase in remote working, rising rates of unemployment, and anxieties and uncertainties for workers around what the ‘new normal’ will be [3].

The pandemic has brought major disruption to the performing arts industry – an industry known for already precarious, often freelance, employment prior to the pandemic [4, 5] with professionals in the industry often unprotected by employment regulations and unable to plan for the future [6]. In early 2021 the Actors Fund, a non-profit organisation supporting the entertainment community in America, surveyed 7,163 creators and actors during the pandemic and found that 76% reported loss of income, 62% lost part-time or gig employment, and 50% lost full-time jobs [7]. The cultural and creative sectors have also been labelled one of the most negatively-affected sectors in Europe [8]. Countries around the world have seen the closure of entertainment venues such as theatres and concert halls [9], thus denying performing artists their traditional communication with the public [10]. In addition to losing work, many performing artists found themselves unable to rehearse with their usual groups due to restrictions on social contact, which could result in performers being under-prepared to re-join the workforce post-pandemic [11].

Psychological wellbeing has been a major concern across the world throughout the pandemic, given the potential detrimental effects of quarantine such as anxiety, depression, loneliness and anger [12] and the impact of prolonged isolation from others on humans who are, at heart, ‘social animals’ [13]. Wellbeing of performing artists may be a particular concern.

Professionals in the performing arts sector often report high levels of organisational demands, poor job security, over-commitment, difficulties coping with critical performance feedback, perceived lack of recognition for their work and poor relationships with colleagues (including competition amongst peers, bullying and temporary relationships due to transient affiliation to organisations), all of which may negatively impact on wellbeing [14]. Additionally, poor psychological wellbeing has been associated with the financial impact of infectious disease outbreaks [12], raising concerns for those who earn even part of their living from performing. Those involved in performing arts who do not earn their living from performing are also likely to have been impacted by the social restrictions of the pandemic, as the social aspect of performing appears to substantially improve psychological wellbeing [13]; the sudden loss of in-person performances may therefore negatively impact performers.

We were therefore interested in exploring how the COVID-19 pandemic has affected performing artists. Our target population was, broadly, ‘individuals for whom performing arts are a central part of their lives’. Such a target population cannot be easily defined by boundaries such as ‘being a professional working in the industry’ or ‘possessing a formal level of education in the performing arts’ because many performing artists do not hold formal qualifications or have an income exclusively reliant on their artistic [16]. The review was therefore interested in, but not limited to, professional performing artists; we were also interested in the experiences of amateur performers and performing arts students and teachers.

The aim of this scoping review was to explore published literature on how those in the performing arts have experienced the pandemic - in particular, how lockdown has affected their wellbeing and the challenges and opportunities they have experienced.

## 2. Materials and method

This review followed Arksey and O’Malley’s scoping review framework [17], consisting of the following stages: identifying the research question; identifying relevant studies; selecting studies for inclusion in the review; charting the data; and collating, summarising and reporting results.

### 2.1. Identifying the Research Question

The following question was identified: What is known about the impact of the COVID-19 pandemic, and its associated lockdowns and restrictions, on the wellbeing of performing artists (including both individuals working in the performing arts sector and non-professionals who are members of performing arts organisations), and what challenges and opportunities have they faced during the pandemic?

### 2.2. Identifying Relevant Studies

The authors designed the following search strategy: (coronavirus or covid* or sars-cov-2 or lockdown* or pandemic*) AND (performing art* or musician* or artist* or dancer* or actor* or actress* or creative art* or creative occupation* or singer* or performer* or entertainer* or entertainment industry) This strategy was used to search six electronic databases (Embase, Global Health, Medline, PsycInfo, Social Policy and Practice, and Web of Science) on November 30^th^ 2021. The reference lists of all studies deemed appropriate for inclusion were also hand-searched.

### 2.3. Selecting Studies for Inclusion

Studies were eligible for inclusion if they:

- Were published in English, as this is the language spoken by the authors;
- Presented results of peer-reviewed research;
- Had a study population of greater than one (i.e. no case studies);
- Had a study population consisting of individuals working in the performing arts sector *or* non-professionals for whom performing arts was a central part of their lives, such as amateur performers or performing arts students;
- Considered the experiences of performing artists during the COVID-19 pandemic in terms of: direct measures of wellbeing such as stress and mental health; indirect measures of wellbeing i.e. other relevant factors which might impact on wellbeing, such as social connections and finances; and any benefits or challenges experienced due to the pandemic-related social restrictions.

Resulting citations were downloaded to EndNote© reference management software (Thomson Reuters, New York). The first author screened the titles of all citations for relevance to the review, excluding any clearly not relevant to the study’s aims. The abstracts of remaining citations were then screened, with any clearly not meeting the inclusion criteria being excluded. Finally, the full texts of remaining citations were obtained and screened to ensure they met all inclusion criteria. A random sample of approximately 10% of initial citations were also screened by the second author to ensure reliability of the screening process; any disagreements were discussed between the authors until consensus was reached.

### 2.4. Charting the Data

Data was extracted onto a Microsoft Excel spreadsheet containing the following headings: first author; year of publication; country of study; methodological design; number of participants; socio-demographic characteristics of participants; measures used; key results; conclusions; and limitations. Thematic analysis [18] was used to group the results of included studies into a typology.

### 2.5. Collating, Summarising and Reporting Results

Each of the themes identified in the data is summarised in the Results section below, with a narrative description of each theme and discussion of the evidence within each theme. More detailed results of each study are presented in Supplementary File 1.

## 3. Results

A total of 1,199 citations were found via the database searches, and 90 duplicates were removed. Of the remaining citations, 1,000 were excluded based on title and 79 were excluded based on abstract. After screening the full texts of remaining citations, a further 10 were excluded and two additional references were added from hand-searching the reference lists of the full texts. One paper which passed the abstract screening was not available in full online and the authors did not respond to a request for the full text, therefore this too was excluded.

The majority of the included studies were from western countries, with nine from Europe (including participants from Belgium, Germany, Italy, the Netherlands, Norway, Sweden, and the United Kingdom (UK)). Two studies were from Australia and one from the United States of America (USA); one additional study included participants from both the USA and Canada, whilst another included participants from Australia, the USA and the UK. Two studies were from Asia (China, n=1 and Israel, n=1) and one was from South America (Brazil). Three studies included participants from multiple countries across the world, and one study did not make it clear where participants were from (although the authors were all based in Europe). Study population sizes ranged from 18 – 5,044 (mean: 568, median: 101). Participants in six studies were students or teachers of performing arts, music or music education. Three studies included professionals working in any area of the performing arts, whilst another three focused on professional musicians. Five studies included professional and/or amateur participants who were members of music groups (such as choirs or orchestras) whereas one study described participants simply as ‘musicians’, one described participants as ‘professional or non-professional voice users’ and two others described participants as ‘music creators’ or ‘producers of art’. Eight studies were qualitative in nature, using either interviews or qualitative open-ended surveys, whilst twelve were quantitative and one used both qualitative and quantitative methods.

An overview of the characteristics of included studies is provided in Table A.1.

**Table A.1.** Characteristics of included studies

| **Authors**<br>**(year)** | **Country** | **Participants** | **Participant ‘n’,**<br>**demographics** | **Measures,**<br>**dates of data**<br>**collection** |
| --- | --- | --- | --- | --- |
| Antonini<br>Philippe et<br>al. (2020)<br>[27] | Authors<br>based in<br>Switzerland,<br>Austria and<br>Italy; unclear<br>where<br>participants<br>were from | Music teachers and<br>their students, who<br>were in their last year<br>of a Master’s degree<br>in music and earned<br>their living from<br>music (vs. athletes<br>who were university<br>students in sports<br>sciences and held<br>national titles, and<br>their coaches) | 18 (6 musicians and<br>3 teachers vs. 6<br>athletes and 3<br>coaches)<br>Mean age of entire<br>sample 33.5<br>Gender breakdown<br>of entire sample:<br>66.7% male, 33.3%<br>female | Semi-structured<br>interviews<br>asking about<br>different<br>aspects of the<br>teacher/student<br>or coach/athlete<br>relationship<br>during<br>lockdown<br>Dates of data<br>collection not<br>reported |
| Cohen &<br>Ginsborg<br>(2021) [22] | UK | Musicians (freelance,<br>self-employed<br>orchestral musicians) | 24<br>Mean age 50.62; 12<br>mid-career | Semi-structured<br>interviews<br>asking about |
|  |  |  | musicians (aged 35-45, mean 40.75); 12 older musicians (aged 53+, mean 60.5)<br>41.7% male, 58.3% female | participants' careers pre-pandemic and the impact of COVID-19 on their lives<br>Data collected May-June 2020 |
| Crosby & McKenzie (2021) [21] | Australia | Professional musicians | 203<br>Mean age not specified; biggest age group was 30-39 (17.2%)<br>59.6% male, 39.9% female, 0.5% not specified | Study-specific survey assessing pre-pandemic employment and income and how these have been affected by the pandemic<br>Data collected June-July 2020 |
| Daffern et al. (2021) [25] | UK | Choir members and facilitators/conductors who participated in virtual choirs during lockdown | 3,948 (819 facilitators; 2,753 choir members)<br>Age not reported<br>Facilitators: 35.8% male, 63.2% female, 0.9% other;<br>Choir members: 21.5% male, 77.9% female, 0.6% other | Study-specific survey assessing use and experiences of virtual choirs<br>Data collected July–August 2020 |
| Davis & Phillips (2021) [37] | Australia, but participants were from Australia (n=56), New Zealand (n=2), Mexico (n=1), Singapore (n=1) and the USA (n=1) | Performing arts teachers or drama teachers | 61<br>Age not reported<br>Gender not reported | Qualitative survey with 16 open-ended questions about how COVID-19 impacted teaching and learning, teacher perceptions about student learning and how teachers coped during the pandemic |
|  |  |  |  | Data collection began in May 2020 and continued throughout 2020 |
| Draper & Dingle (2021) [38] | Australia, USA and UK | Music group members (singing groups, instrumental groups or dance groups) | 257 (172 members of singing groups, 48 members of instrumental groups, 37 members of dance groups)<br>Mean age 45.93<br>22% male, 78% female | Study-specific survey assessing the nature of group membership pre-pandemic; group identification (Doosje et al.'s Group Identification Scale); psychological need satisfaction (adapted version of Kyprianides & Easterbrook's scale); extent to which their music group had adapted to social distancing by meeting electronically; nature and success of this adaptation or reasons for non-adaptation; mental health (mental health score of the SF-12); extraversion (subscale of the Mini-IPIP)<br>Data collected May 2020 |
| Flore et al. (2021) [20] | Australia | Professionals currently working in | 40 | Qualitative survey with 9 |
|  |  | an arts or creative role | Mean age not reported; 15 between 40-49, 8 between 18-29, 10 between 30-39, 7 aged 50+<br>24 female, 12 male, 3 non-binary or gender diverse, 1 did not disclose | open-ended and 9 multiple-choice questions asking about the extent to which participants experienced changes in their work; their use of social and digital media to continue their work; and mental health experiences before and after the restrictions<br>Data collected August-October 2020 |
| Fram et al. (2021) [28] | USA & Canada | Music creators | Surveys: n=101; Interviews: n=12<br>Surveys: Mean age not reported – the majority were between 25-44; Interviews: Age not reported<br>Surveys: 64.4% male, 33.7% female, 2% other<br>Interviews: Gender not reported | Survey: study-specific survey assessing individual musical practice and inspirations, genre and style identification, collaboration network and behaviours, and technology use before the pandemic, during the early stages of the pandemic, and in the most recent month<br>Interviews: semi-structured interviews on the same topics as the survey |
|  |  |  |  | Dates of data collection not reported |
| Frick et al. (2021) [16] | Germany | Producers vs. consumers of art (performative arts, music, other forms of cultural engagement) | 828 (301 producers v 295 consumers v 232 consumer/producers)<br>Mean age not reported; over half of participants were aged over 50<br>50.9% male, 48.2% female, 1.0% gender-diverse | Study-specific survey assessing type and frequency of participation in cultural events in the 30 days leading up to lockdown; type and number of cancelled events; Kemper et al. scale of optimism-pessimism; specific consequences of 'cultural withdrawal' experienced<br>Data collected May–June 2020 |
| Li et al. (2021) [34] | China (Hong Kong) | Teachers and students from a Performing Arts Academy | 76 teachers, 163 students<br>Age not reported<br>Gender not reported | Study-specific survey to assess experiences with hybrid pedagogy, including technology and the e-learning environment<br>Data collected July 2020 |
| MacDonald et al. (2020) [29] | UK (participants were international) | Members of the Glasgow Improvisers Orchestra, which grew to include musicians from across the world during the pandemic | 29<br>Age not reported<br>Gender not reported | Qualitative interviews about experiences performing online with the orchestra<br>Dates of data<br>collection not specifically reported, although interviews took place after three months of virtual improvisation sessions |
| Morgan-Ellis (2021) [36] | USA | Sacred Harp singers (singers who sing from The Sacred Harp, a collection of shape-note songs) | 22<br>Age not reported<br>Gender not reported | Fieldwork and reflections as a participant-observer;<br>Interviews about experiences of online singing during the pandemic<br>Fieldwork data collection began in April 2020 and continued for 3 months;<br>Interview data collected July-September 2020 |
| Onderdijk et al. (2021) [33] | Belgium (participants were from Belgium and the Netherlands) | Musicians | 234<br>Mean age 38.5<br>47% male, 52.6% female. 0.4% preferred not to disclose | Study-specific survey assessing experiences of joint music-making before and during lockdown<br>Data collected April-May 2020 |
| Primov-Fever et al. (2020) [30] | Israel | Professional artistic voice users vs. nonprofessional voice users | 110 (57 professionals, including 34 singers and 19 actors, vs. 53 nonprofessionals) | Voice COVID-19 questionnaire designed specifically for this study, consisting of |
|  |  |  | Mean age 37.72<br>35.5% male, 64.5% female | 20 items designed to assess general concern about voice, current voice difficulties, vocal training, current stress, and vocal hygiene; Singing Voice Handicap Index to assess self-perceived voice concerns; Generalized Anxiety Disorder (GAD-7) to assess anxiety<br>Data collected April 2020 |
| Rosset et al. (2021) [26] | Germany | University students enrolled in performance and music education programmes | 80, including 54 music education majors and 26 performance majors (vs. retrospective data from 75 students at the same university in 2019)<br>Mean age: 20.85 (vs. 21.11)<br>47% male, 53% female (vs. 44% male, 56% female) | University-specific wellbeing and health survey, with added questions relating to COVID-19, perception of university's handling of the pandemic, fear of health problems, and the Stress and Coping Inventory to assess coping strategies<br>Data collected July 2020 (and compared with data collected June-July 2019) |
| Schiavio et al. (2021) [35] | Italy | Music students at an Italian conservatory | 20<br>Mean age 31.5<br>65% male, 35% female | Open-ended questionnaire designed to assess dimensions of musical learning impacted by lockdown measures<br>Dates of data collection not reported |
| Spiro et al. (2020) [19] | UK | Professionals for whom at least part of their livelihood came from working in the arts and cultural areas | Study 1: 385<br>Mean age 44.08<br>63% female, 35% male, 2% non-binary/transgender, 0.3% preferred not to disclose<br>Study 2: 341<br>Mean age 44.41<br>64% female, other percentages not reported | Study 1: HeartS Professional survey charting working patterns, income, sources of support, indicators of mental and social wellbeing; Mental Health Continuum – Short Form to assess mental health; Centre for Epidemiologic Studies Depression Scale to assess depressive symptoms; Social Connectedness Scale to assess connectedness to others; Three-Item Loneliness Scale to assess loneliness<br>Study 2: |
|  |  |  |  | HeartS Professional Survey two open-ended questions on how the pandemic impacted on arts and cultural work and how the pandemic impacted on health and wellbeing<br>Data collected April–June 2020 |
| Stubbe et al. (2021) [31] | Netherlands | Performing arts students | 98 (32 studying dance, 32 studying to be dance teachers, 25 studying circus, 9 studying music theatre)<br>Mean age 19.89<br>24.3% male, 75.7% female | Performing Artist and Athlete Health Monitor to assess mental health, stress, sleep quality and loneliness<br>Data collected monthly between September 2019 – May 2020 |
| Szostak & Sulkowski (2021) [23] | Poland (participants were international) | Professional artists with additional, varying levels of, entrepreneurship | 21 (majority – 86% - had a music background)<br>Age not reported; year of birth ranged from 1947-1999<br>71% male | Qualitative data collected via interview or 36-question survey; unclear which questions were asked<br>Majority of data collected February-March 2020 |
| Teixeira et al. (2021) [24] | Brazil | Music professionals (musicians, public private institutions) | 174<br>Mean age not | Study-specific survey to assess the |
|  |  | and their actions towards the music sector, managers or owners of show/entertainment places) | reported; 36 aged 18-30, 54 aged 31-40, 46 aged 41-50, 35 aged 51+<br>77.6% male, 19% female, 0.6% non-binary, 1.1% preferred not to disclose (unclear why these do not total 100%) | impact of COVID-19 on musical professionals whose income depended mostly on face-to-face activities and how they survived financially<br>Data collected August–October 2020 |
| Theorell et al. (2020) [32] | Sweden & Norway | Members of national choir organisations, both amateur and professional | 3,163 Swedish and 1,881 Norwegian<br>Mean age not reported; 66% were aged between 31-65<br>76.8% female, 23.2% male | Study-specific survey; participants asked to rate the importance of various aspects of choir singing (social bonding, something to look forward to, possibility to experience flow, aesthetic experiences, the physical exercise inherent in singing, breathing training, and voice training) and rank how much they missed them during lockdown<br>Dates of data collection not clear, but the aim of the study was to assess the |
|  |  |  |  | March–May<br>2020 lockdown<br>experience |

Thematic analysis identified seven main themes (loss of work, financial impact, concerns about the future, psychological wellbeing, social connections, continuing creative pursuits during lockdown, and inequalities) and a number of subthemes. These are summarised in Table A.2.

**Table A.2.** Summary of themes and subthemes

| Theme | Description |
| --- | --- |
| Loss of work | • For professionals, pandemic was seen as worsening an already unstable, precarious type of employment<br>• Negative impact on employment activities<br>• Sudden nature of employment cessation experienced as difficult |
| Financial impact<br>*Subtheme: Loss of income*<br>*Subtheme: Financial support*<br>*Subtheme: Impact of financial losses* | • Substantial reduction in income for those whose livelihood depended on performing arts<br>• Many were forced to seek financial support e.g. from government schemes, but often found they were ineligible<br>• Many reported a sense of injustice in the perceived lack of support for performing artists<br>• Participants experienced anxiety and poor wellbeing as a result of financial hardship |
| Concerns about the future<br>*Subtheme: Personal future as a performing artist*<br>*Subtheme: Future of the industry* | • Participants – particularly younger participants – described anxiety and uncertainty around their future careers<br>• Fears for the future of the industry in general |
| Psychological wellbeing<br>*Subtheme: Challenges*<br>*Subtheme: Detrimental effects of the pandemic on psychological wellbeing*<br>*Subtheme: Positive effects of the pandemic on psychological wellbeing*<br>*Subtheme: Factors associated with wellbeing* | • Participants reported a number of detrimental emotional, cognitive-behavioural and health-related effects including anxiety, feelings of powerlessness and disturbed sleep<br>• Wellbeing appeared to be poorer than it was pre-pandemic and also worsened as length of time spent in lockdown increased<br>• However, some participants reported a decrease in stress and burnout, and enjoyed having more free time to spend with family or develop new skills |
|  | Some evidence that being older, male, and partaking in physical exercise were associated with better wellbeing, whilst financial hardship and greater fears of health problems were associated with poorer wellbeing |
| Social connections | Many felt lonely and missed the social component of performing<br>Family and friends not in the performing arts were perceived as not fully understanding the challenges faced, whereas support from other artists was more helpful<br>Being older, taking part in physical exercise, and living with other people were associated with less loneliness and better social connectedness<br>Loneliness worsened as length of time in lockdown increased |
| Continuing creative pursuits during lockdown<br>*Subtheme: Maintaining creative identity*<br>*Subtheme: Seeking new avenues*<br>*Subtheme: Adapting to online platforms*<br>*Subtheme: Barriers to creative pursuits*<br>*Subtheme: Virtual vs. in-person performing* | Creative pursuits seen as central to participants' identities<br>Some participants sought new collaborations or new creative or entrepreneurial avenues during lockdown<br>The majority reported using online platforms to continue rehearsing, collaborating, and performing<br>Virtual rehearsals or performances were praised for being convenient, flexible, allowing collaboration with others across the world, improving confidence to try new things and allowing a sense of belongingness<br>Many barriers to continuing creative pursuits from home, such as lacking motivation and issues with the home environment<br>Many barriers specific to online platforms, such as lack of access to appropriate technology or equipment, internet issues, lack of technical expertise, and 'Zoom fatigue'<br>For many, virtual modes were a poor substitute for in-person performing and could not replace the sound, |
|  | energy and quality of an in-person performance |
| Inequalities | • Racial disparities regarding loss of income, with black performers more severely financially affected than white performers overall<br>• Concerns about how participants with disabilities or special needs could cope with adapting to remote performing<br>• Concerns for disadvantaged groups such as those lacking the finances needed for shifting to remote platforms or those living in unsuitable home environments<br>• Those with caring responsibilities experience additional pressures |

### 3.1. Loss of Work

Employment in the performing arts industry was described as being irregular and unstable prior to the pandemic, with professional participants reporting the COVID-19-related restrictions exacerbated the already precarious nature of their employment [19] and intensified their feelings of job insecurity and instability [20]. Participants working in the industry unsurprisingly reported a loss of employment due to COVID-19 restrictions – in Crosby and McKenzie’s study of professional musicians [21], 99% (200/203) reported a negative impact of the pandemic on their employment activities, and 64% reported unemployment (compared to 13% the previous year); in Spiro et al.’s study [19], 71% of 385 participants reported spending less time working than pre-pandemic. In the latter study, the areas of work with the largest reductions were those which involved predominantly working with others and working offline; 96% reported less time was spent performing, and the majority also reported a reduction in time spent conducting, directing, teaching or mentoring, managing, promoting, composing and choreographing. The pandemic-related loss of work was experienced as being very sudden, with the creative arts industry perceived as shutting down overnight [20, 22] resulting in feelings of grief and loss [20]; this sudden cessation of work was often emotionally overwhelming, particular for younger professionals [22].

### 3.2. Financial Impact

#### 3.2.1. Loss of Income

Given the substantial reduction in work for the majority of professionals working in the performing arts field, it is unsurprising that many also reported that the pandemic had a profound financial impact [19,20,21,22,23,24]. In several studies, over 70% of participants reported income reductions [19,21,24; over half (54%) of Spiro et al.’s [19] 385 participants considered themselves to be in financial hardship whilst almost half (43.9%) of Teixeira et al.’s [24] 174 participants were earning minimum wage or less. Daffern et al.’s [25] choir member and facilitator participants also described a negative economic impact due to loss of funding and personal income, but additionally reported economic advantages to virtual choir participation such as reductions in travel costs and rehearsal venue costs. Although many participants across studies had made adaptations such as shifting to remote work, most professionals were still unable to maintain their usual income due to loss of customers and reduced demand.

#### 3.2.2. Financial Support

Participants in several studies reported applying for income support, grants and government schemes to provide self-employed workers with financial assistance [19,20,21,22]. The number seeking financial support appeared to have significantly increased from previous years: for example, Crosby and McKenzie’s study [21] of 203 professional musicians found an increase in participants seeking government income to 60% from 9% the previous year. However, many found they were rejected as they did not meet the criteria for support [20,21,22], for reasons such as earning over the threshold, recently working abroad, being foreign nationals [22] or already receiving income support [21]. Flore et al.’s [20] participants described the government’s approach to financial support as ‘one-size-fits-all’ and incompatible with the performing arts industry, leaving them feeling misunderstood.

Similarly, several of Cohen and Ginsborg’s [22] musician participants expressed a sense of injustice in the government’s treatment of musicians and perceived lack of support for the arts, and Spiro et al.’s [19] participants reported ‘falling through the cracks’ and described the perceived lack of support from the government as leading them to feel under-valued.

Participants also found themselves relying on partners’ incomes, borrowing money from family or friends, or accessing early superannuation [21] or borrowing money from colleagues [19]. Participants were more reluctant to seek donations for their work from the public, with Flore et al.’s [20] participants reporting feeling that it was inappropriate at a time of global crisis.

#### 3.2.3. Impact of Financial Losses

Participants described feeling anxiety about money [22] and fears that their pre-COVID-19 income would never return [21]; those who experienced financial hardship reported significantly lower wellbeing, greater depressive symptoms and greater feelings of loneliness [19] and poorer mental health generally [26].

### 3.3. Concerns about the Future

Participants across studies described concerns about the future, both their own futures as performing artists and the future of the industry itself.

#### 3.3.1. Personal Future as a Performing Artist

Several of Cohen and Ginsborg’s [22] musician participants described anxiety around their future career and many described difficulties coping with this career uncertainty; younger participants expressed confusion and distress about what they should be doing and several participants were considering leaving the profession and either retraining (younger participants) or retiring early (older participants), or expanding their work to include areas other than performing, such as teaching. Spiro et al.’s [19] participants also frequently reported uncertainty regarding future work and worries about their career progression, whilst Antonini Philippe et al.’s [27] postgraduate music student participants reported feelings of destabilisation and doubt about their future careers. Flore et al.’s [20] participants reported feeling that the uncertainty of the sector was likely to extend beyond the pandemic and felt pessimistic and hopeless about their futures. In Crosby and McKenzie’s study [21], whilst many participants were optimistic about continuing creative employment, they reported low confidence in demand returning for their own artistic services and for live performances in general.

#### 3.3.2. Future of the Industry

Many participants were doubtful about the future of the performing arts industry. Flore et al.’s [20] participants (creative arts professionals) reported feeling the creative arts sector was devalued, misunderstood and ignored even prior to the pandemic, and these feelings were exacerbated by the pandemic. Szostak and Sulkowski’s [23] participants also reported that the pandemic had left them feeling unwanted and unimportant to society. Participants described anxiety about the future of their profession in general and the state of the arts sector [20, 22].

### 3.4. Psychological Wellbeing

#### 3.4.1. Detrimental Effects of the Pandemic on Psychological Wellbeing

Across studies, performing artists reported various detrimental emotional, cognitive-behavioural and health-related effects of the pandemic. These included anxiety and distress [19,20,22,28,29,30]; new or worsening symptoms of depression [19]; mood disturbances [19, 22]; panic [28]; exhaustion or fatigue [19, 29]; concentration difficulties [19]; confusion [29]; boredom [19]; low motivation [19]; self-doubt [20]; feelings of powerlessness [20, 29]; desperation [28]; anger [29]; disconnection from others [20]; guilt at not working and feeling lazy [20]; mentally ‘shutting down’ and detaching to avoid thinking about the loss of performing [29]; fear for the health of themselves or their loved ones [19, 22]; disturbed sleep or insomnia [19,22,29]; and poor eating or drinking habits [19]. Some reported feeling shame about their negative emotions, perceiving that they were unjustified when others were in worse situations [20].

Although most studies were cross-sectional, a minority compared data collected during the pandemic with responses to the same surveys by the same populations in previous years, whilst others asked participants to retrospectively rate their pre-pandemic wellbeing. We therefore have at least some estimates of the extent to which wellbeing was negatively affected by the pandemic: for example, 72% of Crosby and McKenzie’s [21] 203 participants reported they were less satisfied with their lives than they were in the previous year, 85% of Spiro et al.’s [19] 385 participants reported feeling more anxious than pre-pandemic, and Stubbe et al.’s [31] participants reported significantly more mental health complaints than pre-lockdown. Meanwhile, Rosset et al. [26] found that performing arts and music education students reported more stressful thoughts and feelings during the pandemic than before, although there were no significant differences in self-assessed symptoms of anxiety or depression between students who responded during the pandemic and those who had responded to the same surveys a year earlier (pre-pandemic). Additionally, psychological wellbeing appeared to worsen over time during the pandemic, as the length of time spent in lockdown increased [19, 20].

#### 3.4.2. Positive Effects of the Pandemic on Psychological Wellbeing

Across studies, participants also reported various benefits arising from the lockdown period.

In Spiro et al.’s [19] study, 55% (of 385) reported moderate levels of wellbeing, including 34% who reported they were ‘flourishing’ (compared to 11% who felt they were ‘languishing’).

Stubbe et al. [31] noted that average stress scores fell significantly from pre-lockdown to lockdown, and sleep quality improved significantly during lockdown. Participants reported reduced feelings of burnout [20]; less pressure [19]; more time for themselves [19]; more time to spend with family [20]; enhanced connections with others [19] and having time to learn new non-performing-related skills such as learning a new language or doing voluntary work [22]. Lockdown also led to new insights, such as no longer taking things for granted [22]. Some participants reported improvements to their health and wellbeing due to exercising more, drinking less and experiencing less stress [19, 22]. Additionally, participants reported improvements to their performing arts skills, due to more time to rehearse [26]; expanding their skillsets [19,22,28] and realisations about their preferred creative and collaborative processes [28].

#### 3.4.3. Factors Associated with Psychological Wellbeing

Statistical analyses suggested that better psychological wellbeing during the pandemic appeared to be associated with older age [19, 22] with older participants more likely to reframe the experience as temporary, often because they had experienced and overcome other challenging times in their professional lives [22]. Better psychological wellbeing also appeared to be associated with being male [19, 26] and taking part in physical exercise, both prior to and during the pandemic [19]. Those who had a greater general fear of health problems appeared to have significantly poorer mental health than those who did not [26], as did those who experienced financial hardship [19, 26]. Frick et al.’s [16] study found that self-perceived unsatisfactory coping with COVID-19 restrictions was associated with more pre-lockdown cultural events and greater loss of events (i.e. more cancelled events), as well as lower optimism, living in an urban area, and being aged under 60. Some participants self-reported that seeing other members of the arts community struggling worsened their own mental health [20].

### 3.5. Social Connections

Physical restrictions limiting in-person socialising left participants feeling lonely [19] and isolated from their creative communities [20], with the social component of performing reported as the most-missed aspect [32]. The loss of in-person interactions with creative communities had a substantial impact on participants’ wellbeing as such communities were seen as fostering their emotional health [20]. Family and friends not in the creative sector were often perceived as not fully understanding, or minimising, the challenges faced by artists; therefore, support from other artists was particularly valued as they understood the unique challenges faced [20].

Shifting relationships online posed challenges for teachers and students of performing arts, with the need to develop and negotiate new boundaries, and students reported that relationships felt more detached and their teachers were perceived as distant [27].

Spiro et al.’s [19] regression analyses showed that older participants reported significantly more social connectedness and less loneliness, as did participants who took part in physical exercise (both pre-pandemic and during the pandemic). Higher social connectedness and lower loneliness were also associated with earlier completion of the survey (within the first two weeks), suggesting that these might worsen over time during the lockdown period.

Participants who lived alone also reported significantly higher loneliness.

### 3.6. Continuing Creative Pursuits during Lockdown

#### 3.6.1. Maintaining Creative Identity

For professionals in the sector, art, creativity, and working in the arts were seen as central to participants’ identities [20,22,23] and perceived to be a lifestyle, rather than just a job [20]. The reduction in creative activities during lockdown therefore led to a loss of self-worth and loss of a sense of identity [19, 22]. Consequently, many participants reported feeling it was important to continue carrying out creative activities during the pandemic as a way of reconnecting with their identities [22]. Non-professional performing artists expressed similar sentiments, reporting that performing was central to their identities and not being able to perform resulted in feelings of grief and distress [29] and suggesting they needed to continue their creative pursuits to retain a sense of purpose and normality [25]. Continuing to pursue creative avenues with others during lockdown was also a way of feeling connected to others, coping with the stressful situation, and relieving boredom [33].

#### 3.6.2. Seeking New Avenues

The majority of Szostak and Sulkowski’s [23] participants focused on finding new creative and entrepreneurial avenues, leading the authors to conclude that entrepreneurship was one potential way of improving wellbeing during the pandemic. Most of Fram et al.’s [28] participants also reported embarking on new collaborations during the pandemic, although many referred to an initial period of stagnation at the beginning of lockdown followed by an attempt to reclaim old collaborative practice and find new ones.

#### 3.6.3. Adapting to Online Platforms

Most performing artists reported adapting their work to a virtual setting, such as online collaborative rehearsals and recordings or virtual concerts, carried out on platforms such as Zoom. Meanwhile, teachers and students in the performing arts adapted to online learning [27, 34]. Some participants reported that online participation in rehearsals and performances was more convenient, more flexible, less time-consuming and cheaper than in-person [25] whilst students praised online learning for greater flexibility [34, 35], less commuting time [35] and promoting their personal growth and creativity [34, 35]. Virtual interactions were also praised for allowing contact across large distances – for example, allowing collaborations with people in other countries which would not have happened otherwise [28, 36]. The online creative space was praised for being a non-judgmental space for allowing new ways of thinking and unexplored interdisciplinary expressions and choices [29], and some participants reported they had more confidence to try new things and take risks within the space of virtual platforms than they would in person [29, 36]. Virtual participation was viewed as benefiting mental health, allowing participants a sense of purpose, giving them something to focus on, providing a sense of security, belongingness and connection to others, and helping participants to creatively evolve and redefine their identities [29].

#### 3.6.4. Barriers to Creative Pursuits

However, working remotely could also be frustrating. Across studies, participants reported various barriers to continuing their creative pursuits. Some reported finding it difficult to motivate themselves [19,22,26] for various reasons including feeling that it was pointless, being too busy with childcare, and finding it difficult to play alone rather than as part of an ensemble [22]. Participants also reported lacking access to rehearsal rooms, difficulties concentrating, and mental health problems which prevented them from their pursuits [26]. Some reported that additional effort and preparation time was needed in order to participate in remote performances [25]. The home environment also appeared to be a hindrance, with participants citing issues regarding difficulties finding a way to participate without distractions [25]); not being able to sing at full volume [36]; limited space [25, 34] and hard floors making it difficult to dance [34]. Not being able to continue creative pursuits to the extent they wanted left participants feeling their creativity was stagnating and stifled [19]. Performing arts students reported problems with remote learning including teachers having difficulties conforming to new teaching modalities, difficulties showing visual examples when remote teaching/learning, and less feedback from teachers than they would get face-to-face [35]. Teachers cited challenges such as less interaction with students, more theory and less physical work, less time to invest on disruptive students, and some students failing to engage in remote learning [37].

Amateur and professional participants alike also reported barriers specific to virtual collaborating, learning and performing, such as lack of technological expertise and difficulties learning to use remote platforms [24,25,28,33,38]; lack of appropriate technology [25,33,34,35,38]; difficulty organising and co-ordinating a large group of people [38]; lacking the finances needed to invest in equipment to transition to an electronic platform [25, 38]; poor sound quality [36]; and problems with internet connection, bandwidth or Wi-Fi accessibility [25,34,35,36]. Some non-professional participants who were also remotely working other jobs reported ‘Zoom fatigue’ and a reluctance to engage with more screen-based activities [25, 36]. In one study, participants also raised concerns about privacy and security when using Zoom [34]. Onderdijk et al.’s [33] participants reported that non-real-time remote methods (such as playing along with pre-recorded material or recording parts separately from others) had fewer latency issues, and although some were critical of the lack of eye contact and inability to read body language of others, such methods were still reported to lead to a feeling of social connection overall. Rather than highlighting the drawbacks of performing via Zoom, MacDonald et al.’s [29] participants spoke about working with the limitations (e.g. poor internet connection, editing out of sounds), embracing the faults of technology and seeing it as part of their journey.

#### 3.6.5. Virtual vs. In-person Performing

One recurrent theme was that virtual modes of performing were viewed as a poor substitute for in-person performances and the experiences created online did not equal those established offline [22,25,33,34,35,36,38]. In-person performing tended to be viewed as psychologically uplifting, reducing stress and increasing energy, which could not be replaced by online participation [25, 37], which was perceived as more artificial [35]. Virtual performing was reported to lack what Morgan-Ellis’s [36] participants referred to as the ‘surround sound’ of performing with others in person as well as the energy of face-to-face groups [38].

Participants reported they could not get the same quality or ‘feel’ for a performance as they would in a studio [34] and, for choir members, virtual choirs were seen as unable to recreate the real-life experience of voices blending together as well as the social and emotional connection arising from singing together in person [25]. Similarly, performing arts teachers reported they could not replicate online the interplay of energy given and received in the physical presence of one another [37]. Remote rehearsals also left participants feeling they were not developing their skills to the extent they would in real life [25]. For some, virtual participation affected them negatively and created more stress, anxiety, sadness and loneliness, as virtual participation reminded them how much of the social aspect of participation they were missing [25, 36].

### 3.7. Inequalities

One study reported on racial disparities with regard to the financial impact of the pandemic. Teixeira et al.’s [24] study on the impact of the pandemic on the income of 174 professional musicians found that, pre-pandemic, black participants already had a monthly income lower than those who identified as white; during the pandemic, white professionals represented 91.7% of the highest income range whereas black participants, although representing only 37.4% of total respondents, represented 50% among the lowest income range and only 7.7% of those with the highest income.

In Davis and Philipps’ [37] study of performing arts teachers, participants reported extra concern for students with disabilities or learning needs and how such students would adapt to online learning. They also expressed concern for equity and inclusion for disadvantaged groups, for example those students who did not have access to the technology and tools required for online learning, or those with other environmental factors in the home preventing them from engaging meaningfully with their work. Additionally, a number of participants in two other studies reported experiencing further stress and strain due to caring responsibilities or home-schooling children [19, 22].

## 4. Discussion

Our scoping review identified numerous personal and professional challenges and opportunities faced by performing artists during the COVID-19 pandemic. Unsurprisingly, participants reported substantial losses in work and income. Participants in several studies described the perceived failure of governments to understand the nature of work in the industry, reporting ‘falling through the gaps’ when it came to financial support schemes. This supports anecdotal evidence that initial policy responses in Europe were inappropriate, insufficient, and involved problematic assumptions about how performing artists can be productive in a global pandemic [6]. It is important to note that the majority of the data reviewed in this article was collected early on in the pandemic, in the first few months of 2020. Since this time, policy responses have changed in order to promote recovery in the performing arts industry – for example in Australia, several initiatives have been set up to assist such industries including grants, resilience funds and a series of webinars for artists to offer support and advice to those feeling isolated [21]. In Canada, funding programmes have been launched to help the country’s arts community shift their work to digital platforms for online audiences [39]. Various charities and non-profit organisations in the USA have also provided emergency relief for artists [40]. However, whilst such schemes may address short-term financial concerns they will likely do little to help concerns about the future and about employment in the long term.

The loss of work and income raises concerns about the psychological wellbeing of performing artists at such a time, given that financial difficulties due to pandemic-related quarantine have been shown to be associated with poorer mental health even during the relatively short quarantine periods during earlier pandemics, such as the severe acute respiratory syndrome (SARS) pandemic [12]. The unprecedented length of lockdowns and social restrictions during the COVID-19 pandemic has likely led to far greater financial distress and also potentially greater emotional distress. Indeed, psychological wellbeing was reported to have been affected in detrimental ways in the studies included in this review, such as increased anxiety, negative emotions, cognitive difficulties such as poor concentration, and poor sleep.

However, participants across studies also reported psychological wellbeing had been affected in positive ways, too. In fact, some reported improvements in their health and wellbeing which they commonly attributed to a reduction in stress, pressure and burnout. This suggests performing artists are a resilient population, and may be indicative of post-traumatic growth, which refers to growth and positive emotional responses as a result of experiencing great adversity [41]. This resiliency is perhaps unsurprising as professionals are familiar with coping with job insecurity and the uncertainty embedded in the nature of the industry (albeit on a smaller scale than a global pandemic); additionally, performing artists are by nature a creative group, and so it is unsurprising that the majority have been able to seek new avenues or adapt their art to other platforms. Further investigation will be needed to explore the levels of, and factors related to, the post-traumatic growth being seen among the performing arts population since the start of the COVID-19 pandemic.

The loneliness, isolation and loss of social connectedness reported by many participants illustrates the importance of having adequate social support. ‘Social support’, at a time of social restrictions, may appear to be a contradiction but is in fact essential to the wellbeing of performing artists (and, indeed, everyone else) during times of stress and uncertainty [12, 42] with social networks and relationships being a key aspect of resilience [43]. In addition, Flore et al.’s [20] finding that many participants reported feeling their challenges were misunderstood and minimised by family and friends not involved in performing arts highlights the importance of performing artists receiving social support from other artists. This may be particularly pertinent for younger artists, who appeared to suffer from poorer wellbeing during the pandemic and may need additional support. Cohen and Ginsborg [22] suggest that seasoned performers are best placed to help younger, less experienced performers through this period of uncertainty, and suggest virtual meetings held by performers’ organisations could help link up performers to support each other.

As performing was such a central part of participants’ identities, the majority sought ways of continuing their creative pursuits throughout lockdown, which for many was reported to help them cope with the pandemic. This is unsurprising as research suggests that engaging in creative pursuits during the pandemic is associated with better psychological wellbeing in the general population [44]; this is arguably even more important for individuals for whom creative pursuits were already important. Creative pursuits during lockdown were continued either by seeking new avenues or adapting their existing creative work to online platforms.

Such adaptations indicate versatility within the population; however, many reported difficulties adjusting to different ways of performing. Participants reported mixed feelings about the adaptation to remote performing and how well this worked as a substitute for in-person performing. The shift to remote platforms was praised for giving participants a sense of purpose and belongingness and a way to maintain their skills; erasing geographical barriers to participation and allowing people across the world to collaborate together; creating new opportunities for experimentation and growth; and helping to develop confidence. However, many missed performing for an audience as well as the shared physical experience of performing with others, and overall it appeared that remote performing was perceived as not being able to capture the unique energy and dynamics of in-person performances. This supports Datta’s [45] suggestion that ‘virtual choirs’ cannot replicate aspects of live performance such as responding to other singers’ breathing and emotional states, and emphasises that remote performances cannot synthesise place, time, affect and emotion. Additionally, many reported barriers to participation including technology issues and problems with the internet and Wi-Fi connections. Whilst similar issues have likely affected people across all sectors working from home during the pandemic, this may be a particularly pertinent issue for performing artists collaborating remotely, where one person with unreliable internet or poor sound quality could significantly affect the entire group.

A small but notable minority of studies included in this review identified issues around inequalities, raising concerns about the potential impact of the pandemic on particular groups including black performers, performers with disabilities or special needs, performers lacking the finances to afford the technology and equipment required for shifting to remote platforms, and performers with caring responsibilities. Similar concerns about additional pressures experienced by disabled artists, artists from working class and diverse communities, and artists who have caring responsibilities were raised in the early days of the pandemic [6], with concerns the pandemic could result in a step backwards regarding improving inequalities in the performing arts.

Overall, our review supported the results of a UK-specific meta-analysis of unpublished results of surveys being carried out by the creative and cultural industry [4]. This analysis revealed that performing artists reported significant reductions in organisational activities and individual workloads, corresponding with a loss of income/revenue; concerns about the long-term survival of organisations or their ability to continue working in the industry; and issues relating to eligibility criteria for government schemes.

There are a number of implications of this review. First, it is important to build networks of performing artists so that they may benefit from supporting others and having the support of others who can relate to their circumstances. Spiro et al. [19] suggest that such networks could also work to identify the challenges faced by artists, come up with solutions to them, and lobby for additional support for those in the arts. Peer support groups have been found to be helpful in strengthening mental health during the pandemic and helping people to cope [46]. During a global pandemic, such networks would be forced to operate mostly remotely, but it is important that those without computers or reliable internet access are not forgotten [47]. As such, we recommend that performing arts networks should aim to put people in contact with others who live close to them who may be able to provide in-person support by joining that person’s social distancing bubble. Telephone peer support may also be useful, as well as potentially helplines specifically for artists to turn to for support. Such helplines already exist in some countries: for example, in 2017 Australia set up the Support Act Wellbeing Helpline available to anyone in Australia who works in the performing arts or creative industries [48] whilst in 2018 the UK launched a 24-hour Theatre Helpline for theatre professionals [49]. It is important that performing artists are made aware of the existence of support tools like these, and also recommended that countries without similar helplines should consider developing them. We would also like to emphasise the importance of such helplines and peer support networks providing support for non-professional performing artists, as well as professionals; whilst professionals will have unique challenges relating to employment and income, amateurs are likely to be experiencing similar psychological effects to the loss of in-person performances and also need support.

Our findings on the barriers to remote performing suggest changes are needed in order for artists to be able to fully engage with online platforms. Since participants overwhelmingly reported that virtual performing was no substitute for the energy of an in-person performance, changes in technology itself are needed in order to capture the group experience better.

Daffern et al. [25] suggest that future technological developments should focus on immersive audio techniques with narrow latency limits in order to better bridge the gap between remote and in-person performing. Additionally, it is important that performing artists are equipped with the skills needed to adapt their work to remote platforms when necessary. Education and training in performing arts should therefore incorporate more on digital technologies and appropriate skills. It is important to note that, despite participants in this review describing issues around using digital technology for their creative pursuits, they did at least have access to such technology in order to try it. Performing artists without the means to attempt remote performing would be unlikely to have participated in the studies reviewed, which predominantly collected data online. There may therefore be a hidden population of performing artists without access to the internet whose experiences of the pandemic are yet to be explored. This highlights what Baker et al. [50] refer to as the ‘digital divide’: the gap between those in society with full access to computers and those without.

Even if allocated resources and support were amply supplied during the pandemic, it is important to address inequities in the performing arts industry which appeared to exist pre-COVID-19 and increased during the pandemic. This review has raised concerns about the particular impact of COVID-19 on performers who are BIPOC (Black, Indigenous and People of Colour), for example. Pre-pandemic reports suggest large funding disparities between predominantly white and BIPOC performing institutions – for example, the Asian American Performers Action Coalition (AAPAC) found that in 2018-2019, 92.2% of government funding for theatre companies (almost $150 million) went to predominantly white institutions whilst theatres of colour received just $12.5 million [51]. Whilst only one study in the review considered race as a factor affecting pandemic experiences, this study found that race was a significant predictor of the financial impact of COVID-19. Similarly, a survey by the Actors Fund and reported in Variety [7] found that BIPOC respondents were more likely to experience food insecurity, housing changes, increased debt and changed utility usage compared to white respondents. This review also raised concerns about those with disabilities, low income, or caring responsibilities, who may find it more difficult to adapt their performing during the pandemic and who are likely to have experienced additional pressures and challenges during lockdown. A key priority for policy-makers across the world when considering how to support those in the arts should be ensuring that the diverse and unique needs of different communities are met.

### 4.1. Limitations

Due to the timing of this review (conducted in November 2021), there was a lack of longitudinal work to be reviewed, with all data in the included studies collected in 2020 and most gathered early in the pandemic when social restrictions had only been in place for weeks or months. However, the review provides a useful snapshot of how performing artists experienced the pandemic-related lockdowns at their most severe. The next step for researchers in the field should be to consider the long-term effects of the pandemic on performing artists.

Due to COVID-19 restrictions, the majority of studies in this review were advertised and recruited for online. This is understandable given the context of the research, but automatically excludes those without internet access and those lacking the skills and technology to respond to online surveys or take part in remote interviews – which is problematic, given the concerns discussed about additional pressures of the pandemic for disadvantaged groups. Additionally, due to the fact that participation in most studies involved being online, samples were likely to be skewed toward those who had adapted at least somewhat to remote performing, and results may therefore not accurately capture the barriers experienced by others who had not. Many studies also had small sample sizes and were based on convenience samples, meaning results may not be generalisable to the wider performing arts population.

In terms of our own review process, one limitation is the decision to limit to English-language papers; although the included studies represented a number of different countries, important papers in other languages may have been missed. Using more terms in the search strategy may also have yielded additional papers, as could searching more than six databases or including grey literature.

## 5. Conclusions

The COVID-19 pandemic and associated lockdowns and social restrictions have led to substantial losses in work and income for many performing artists and led participants to worry about their futures and that of the industry in general; additionally, they have had to deal with feelings of loneliness and isolation due to social restrictions. Whilst many negative psychological effects of lockdown were experienced, including anxiety and poor sleep, participants also reported positive effects, and described personal and professional opportunities the pandemic had brought, such as experiencing less pressure, enjoyment of having time to spend with family, and time to spend pursuing new avenues, developing new skills or improving existing skills. Although performing artists frequently adapted their work – for example, to online platforms – they reported mixed feelings about how well this worked as a substitute for in-person performing and believed that it could not replace the shared experience of performing in person with others. Additionally, many reported barriers to participation including internet connection issues, lack of appropriate technological skills and difficulties finding the time or space to participate from home. Overall, performing artists showed resilience and versatility throughout the difficulties they faced as a result of the pandemic. However, there are a number of inequalities which raise concern and require further research, such as the unique impact of the pandemic on black performers, disabled performers, those with low income who cannot access the technology needed for remote performing, and those with additional caring responsibilities. It is important to build networks of performing artists who can support each other during difficult times, and incorporate more skills and training in digital technologies into performing arts education.

## Declarations

### Disclaimer

SKB is funded by the National Institute for Health Research Health Protection Research Unit (NIHR HPRU) in Emergency Preparedness and Response at King’s College London in partnership with Public Health England (PHE), in collaboration with the University of East Anglia. Sonny S Patel was supported by the Fogarty International Center and National Institute of Mental Health, of the National Institutes of Health under Award Number D43 TW010543. The views expressed are those of the author(s) and not necessarily those of the NHS, the NIHR, the Department of Health and Social Care, the UK Health Security Agency, National Institutes of Health, or any other institution. The funding source had no involvement in study design; collection, analysis or interpretation of data; writing the report; or the decision to submit the article for publication.

### Ethics Approval

Not required as the paper reviews only data already in the public domain.

### Informed Consent Statement

Not applicable – no primary data was collected.

### Author Contributions

**SKB:** Conceptualization, Methodology, Formal Analysis, Investigation, Data Curation, Writing – Original Draft, Visualization, Funding Acquisition **SSP:** Investigation, Validation, Writing – Review & Editing, Visualization

### Data Availability

The datasets generated during and/or analysed during the current study are available from the corresponding author on reasonable request.

## Data Availability

All data produced in the present study are available upon reasonable request to the authors.

## References

1. World Health Organization, WHO Director-General’s opening remarks at the media briefing on COVID-19 – 11 March 2020. https://www.who.int/director-general/speeches/detail/who-director-general-s-opening-remarks-at-the-media-briefing-on-covid-1911-march-2020, 2020 (accessed 7 December 2021).

2. International Labour Organization, ILO Monitor: COVID-19 and the world of work. Second edition. Updated estimates and analysis. https://www.ilo.org/wcmsp5/groups/public/---dgreports/---dcomm/documents/briefingnote/wcms_740877.pdf, 2020 (accessed 7 December 2021).

3. Fouad, N.A, Editor in chief’s introduction to essays on the impact of COVID-19 on work and workers. J Vocat Behav. 119 (2020) 103441.

4. Comunian, R., England, L., Creative and cultural work without filters: Covid-19 and exposed precarity in the creative economy. Cult Trends. 29(2) (2020) 112–128.

5. Hancock, P., Tyler, M., Godiva, M. Thursday night and a sing-along ‘sung alone’: The experiences of a self-employed performer during the pandemic. Work Employ Soc. (2021) doi:10.1177/09500170211045830.

6. Tsioulakis, I., FitzGibbon, A., Performing artists in the age of COVID-19: A moment of urgent action and potential change. http://qpol.qub.ac.uk/performing-artists-in-the-age-of-covid-19/, 2020 (accessed 8 December 2021).

7. Malkin, M., Actors Fund study details devastating impact of COVID-19 on entertainment industry. https://variety.com/2021/film/news/actors-fund-covid-impact-survey-1234967568/, 2021 (accessed 22 December 2021).

8. IDEA Consult, Goethe-Institut, Amann S., Heinsius J., Research for CULT Committee – Cultural and creative sectors in post-Covid-19 Europe: crisis effects and policy recommendations, European Parliament, Policy Department for Structural and Cohesion Policies, Brussels. https://www.europarl.europa.eu/RegData/etudes/STUD/2021/652242/IPOL_STU(2021)652242_EN.pdf, 2021 (accessed 9 September 2021).

9. Moon, S. Effects of COVID-19 on the entertainment industry. IDOSR-JES 5(1) (2020) 8-12.

10. Bronner, S. Here’s to our community. Med Probl Perform Art. 35(4) (2020) 233–234.

11. Stuckey, M., Richard, V., Decker, A., Aubertin, P., Kriellaars, D. Supporting holistic wellbeing for performing artists during the COVID-19 pandemic and recovery: Study protocol. Front Psychol. 12 (2021) 577882.

12. Brooks, S.K., Webster, R., Smith, L., Woodland, L., Wessely, S., Greenberg, N., Rubin, G.J. The psychological impact of quarantine and how to reduce it: Rapid review of the evidence. The Lancet, 395(10227) (2020) 912–920.

13. Pietrabissa, G., Simpson, S.G. Psychological consequences of social isolation during COVID-19 outbreak. Front Psychol. 11 (2020) 2201.

14. Willis, S., Neil, R., Mellick, M.C., Wasley, D. The relationship between occupational demands and well-being of performing artists: A systematic review. Front Psychol. 10 (2019) 393.

15. Gick, M.I. Singing, health and well-being: a health psychologist’s review. Psychomusicology. 21 (2011) 176–207. doi:10.1037/h0094011

16. Frick, U., Tallon, M., Gotthardt, K., Seitz, M., Rakoczy, K. Cultural withdrawal during COVID-19 lockdown: Impact in a sample of 828 artists and recipients of highbrow culture in Germany. Psychol Aesthet Create Arts. 2021 doi:10.1037/aca0000389

17. Arksey, H., O’Malley, L. Scoping studies: towards a methodological framework. Int J Soc Res Methodol. 8(1) (2005) 19–32. https://doi.org/10.1080/1364557032000119616

18. Braun, V., Clarke, V. Using thematic analysis in psychology. Qual Res Psychol. 3(2) (2006) 77–101, doi: 10.1191/1478088706qp063oa

19. Spiro, N., Perkins, R., Kaye, S., Tymoszuk, U., Mason-Bertrand, A., Cossette, I., Glasser, S., Williamon, A. The effects of COVID-19 lockdown 1.0 on working patterns, income, and wellbeing among performing arts professionals in the United Kingdom (April-June 2020). Front Psychol. 11 (2020) 594086.

20. Flore, J., Hendry, N.A., Gaylor, A. Creative arts workers during the Covid-19 pandemic: Social imaginaries in lockdown. J Sociol. 2021 doi:10.1177/14407833211036757

21. Crosby, P., McKenzie, J. Survey evidence on the impact of COVID-19 on Australian musicians and implications for policy. Int J Cult Policy. 2021 doi:10.1080/10286632.2021.1916004

22. Cohen, S., Ginsborg, J. The experiences of mid-career and seasoned orchestral musicians in the UK during the first COVID-19 lockdown. Front Psychol. 12 (2021) 645967.

23. Szostak, M., Sulkowski, L. Identity crisis of artist during the Covid-19 pandemic and shift towards entrepreneurship. Entrepreneurial Bus Econ Rev. 9(3) (2021) 87–102.

24. Teixeira, N., Vianna, G.M., Lima, R., Jauregui, C., Furtado, L., Alberto, T.P., Medeiros, R. Covid-19 impact on the music sector in Belo Horizonte (Minas Gerais, Brazil). Front Sociol. 6 (2021) 643344.

25. Daffern, H., Balmer, K., Brereton, J. Singing together, yet apart: The experience of UK choir members and facilitators during the Covid-19 pandemic. Front Psychol. 12 (2021) 624474.

26. Rosset, M., Baumann, E., Altenmuller, E. Studying music during the coronavirus pandemic: Conditions of studying and health-related challenges. Front Psychol. 12 (2021) 651393.

27. Antonini Philippe, R., Schiavio, A., Biasutti, M. Adaptation and destabilization of interpersonal relationships in sport and music during the Covid-19 lockdown. Heliyon. 6(10) (2020) e05212.

28. Fram, N.R., Goudarzi, V., Terasawa, H., Berger, J. Collaborating in isolation: Assessing the effects of the Covid-19 pandemic on patterns of collaborative behavior among working musicians. Front Psychol. 12 (2021) 674246.

29. MacDonald, R., Burke, R., De Nora, T., Donohue, M.S., Birrell, R. Our virtual tribe: Sustaining and enhancing community via online music improvisation. Front Psychol. 11 (2020) 623640.

30. Primov-Fever, A., Roziner, I., Amir, O. Songbirds must sing: How artistic voice users perceive their voice in times of COVID-19. J Voice. 2020 doi:10.1016/j.voice.2020.07.030

31. Stubbe, J.H., Tiemens, A., Keizer-Hulsebosch, S.C., Steemers, S., van Winden, D., Buiten, M., Richardson, A., van Rijn, R.M. Prevalence of mental health complaints among performing arts students is associated with COVID-19 preventive measures. Front Psychol. 12 (2021) 676587.

32. Theorell, T., Kowalski, J., Theorell, A.M.L., Horwitz, E.B. Choir singers without rehearsals and concerts? A questionnaire study on perceived losses from restricting choral singing during the Covid-19 pandemic. J Voice. 2020 doi:10.1016/j.jvoice.2020.11.006

33. Onderdijk, K.E., Acar, F., Van Dyck, E. Impact of lockdown measures on joint music making: Playing online and physically together. Front Psychol. 12 (2021) 642713.

34. Li, Q., Li, Z., Han, J. A hybrid learning pedagogy for surmounting the challenges of the COVID-19 pandemic in the performing arts education. Educ Inf Technol. 2021 doi:10.1007/s10639-021-10612-1

35. Schiavio, A., Biasutti, M., Antonini Philippe, R. Creative pedagogies in the time of pandemic: a case study with conservatory students. Music Educ Res. 23(2) (2021) 167–178.

36. Morgan-Ellis, E.M. “Like pieces in a puzzle”: Online Sacred Harp singing during the COVID-19 pandemic. Front Psychol. 12 (2021) 627038.

37. Davis, S., Phillips. L.G. Teaching during COVID 19 times – The experiences of drama and performing arts teachers and the human dimensions of learning. Drama Australia Journal. 44(2) (2021) 66–87.

38. Draper, G., Dingle, G.A. “It’s not the same”: A comparison of the psychological needs satisfied by musical group activities in face to face and virtual modes. Front Psychol. 12 (2021) 646292.

39. Risk, L. Imperfections and intimacies: Trebling effects and the improvisational aesthetics of pandemic-era livestreaming. Critical Studies in Improvisation. 14(1) (2021) 1–22.

40. Artwork Archive, Financial relief resources for artists during COVID-19. https://www.artworkarchive.com/blog/financial-relief-resources-for-artists-during-covid-19, 2020 (accessed 22 December 2021)

41. Tedeschi, R.G., Calhoun, L.G. The posttraumatic growth inventory: measuring the positive legacy of trauma. J Trauma Stress. 9 (1996) 455–471.

42. Patel, S.S., Clark-Ginsberg, A. Incorporating issues of elderly loneliness into the coronavirus disease-2019 public health response. Disaster Med Public Health Prep. 14(3) (2020) E13–14. doi:10.1017/dmp.2020.145

43. Patel, S.S., Rogers, M.B., Amlot, R., Rubin, G.J. What do we mean by ‘community resilience’? A systematic literature review of how it is defined in the literature. PLoS Currents, 9 (2017) ecurrents.dis.db775aff25efc5ac4f0660ad9c9f7db2. doi:10.1371/currents.dis.db775aff25efc5ac4f0660ad9c9f7db2

44. Morse, K.F., Fine, P.A., Friedlander, K.J. Creativity and leisure during COVID-19: Examining the relationship between leisure activities, motivations, and psychological well-being. Front Psychol. 12 (2021) 609967.

45. Datta, A. ‘Virtual choirs’ and the simulation of live performance under lockdown. Soc Anthropol. 28(2) (2020) 249–250.

46. Suresh, R., Alam, A., Karkossa, Z. Using peer support to strengthen mental health during the COVID-19 pandemic: A review. Front Psychiatry. 12 (2021) 1119.

47. Conroy, K.M., Krishnan, S., Mittelstaedt, S., Patel, S.S. Technological advancements to address elderly loneliness: Practical considerations and community resilience implications for COVID-19 pandemic. Work Older People. 24(4) (2020) 257–264. doi:10.1108/wwop-07-2020-0036

48. The Arts Wellbeing Collective, Support Act Wellbeing Helpline. https://artswellbeingcollective.com.au/resources/support-act-wellbeing-helpline/, 2021 (accessed 9 December 2021).

49. Musicians Union, 24-hour helpline for theatre professionals launched. https://musiciansunion.org.uk/news/24-hour-helpline-for-theatre-professionals-launched, 2018 (accessed 9 December 2021).

50. Baker, C., Hutton, G., Christie, L., Wright, S., COVID-19 and the digital divide. https://post.parliament.uk/covid-19-and-the-digital-divide/, 2020 (accessed 9 December 2021).

51. AAPAC, The visibility report: Racial representation on NYC stages. http://www.aapacnyc.org/uploads/1/3/5/7/135720209/aapac_report_2018-2019_final.pdf, 2021 (accessed 22 December 2021).

